# Genomic epidemiology of the cholera outbreak in Malawi 2022-2023

**DOI:** 10.1101/2023.08.22.23294324

**Authors:** Lucious Chabuka, Wonderful T. Choga, Carla N. Mavian, Monika Moir, Houriiyah Tegally, Eduan Wilkinson, Yeshnee Naidoo, Rhys Inward, Christian Morgenstern, Samir Bhatt, G. R. WilliamWint, Kamran Khan, Isaac I. Bogoch, Moritz U.G. Kraemer, Cheryl Baxter, Massimiliano Tagliamonte, Marco Salemi, Richard Lessells, Collins Mitambo, Ronald Chitatanga, Joseph Bitilinyu- Bango, Mabvuto Chiwaula, Yollamu Chavula, Mphatso Bukhu, Happy Manda, Moses Chitenje, Innocent Malolo, Alex Mwanyongo, Dr Bernard Mvula, Dr Mirrium Nyenje, Tulio de Oliveira, Dr Mathew Kagoli

**Affiliations:** Public Health Institute of Malawi (PHIM), Ministry of Health, Malawi; Centre for Epidemic Control and Innovation (CERI), School of Data Science and Computational Thinking, Stellenbosch University, Stellenbosch, South Africa; Emerging Pathogens Institute, Department of Pathology, College of Medicine, University of Florida, United States of America; KwaZulu-Natal Research Innovation and Sequencing Platform, University of KwaZulu-Natal, Durban, South Africa; Department of Biology, University of Oxford, United Kingdom; Imperial College London, United Kingdom; Cophenhagen University, Denmark; BlueDot, Toronto, Canada; Department of Medicine, Division of Infectious Diseases, University of Toronto, Canada; Pandemic Sciences Institute, University of Oxford, United Kingdom; Environmental Research Group Oxford Ltd, c/o Department of Biology, University of Oxford, United Kingdom

**Author notes:** these authors contributed equally (Lucious Chabuka, Wonderful T. Choga, Carla N. Mavian, Monika Moir).

## Abstract

Since early 2022, in the aftermath of two extreme weather events, Malawi experienced its largest ever cholera outbreak, with over 58,000 reported cases and 1,761 deaths as of May 2023. We generated 49 high-quality, near-complete Vibrio cholerae genomes in Malawi from isolates collected between December 2022 and March 2023 from all three regions of Malawi. Using phylogenetic methods with 2159 publicly available genomes, we present evidence suggesting that the Malawi outbreak strains originated from the Pakistan outbreak, the estimated most recent ancestor of this lineage, named T15, was during the Pakistan floods, and once introduced into Malawi was exacerbated by major floods between June and October 2022. The extreme weather events and humanitarian crises in Malawi provided the environment for the spread of Vibrio cholerae, and the subsequent movement of large numbers of people may have facilitated its spread to susceptible populations in areas relatively unaffected by cholera for over a decade.

## Introduction

Cholera is an acute diarrhoeal disease caused by ingestion of food or water contaminated with the bacterium *Vibrio cholerae*^1^. Since mid-2021, there has been an acute upsurge of the seventh cholera pandemic associated with the *V. cholerae* O1 El Tor biotype. Several large outbreaks have occurred in endemic and non-endemic countries, with African countries particularly heavily affected^2^. These outbreaks have been driven by multiple factors, including extreme weather events, humanitarian crises, overlapping health emergencies (particularly with the COVID-19 pandemic) and consequently overstretched health systems^3,4^.

Cholera was first reported in Malawi in 1973 and has been endemic since 1998 with annual increases in incidence occurring during the rainy season (November-May), particularly in the southern part of the country^5^. Prior to 2022, Malawi had experienced three large cholera outbreaks: in 1998-1999, 2001-2002, and 2008-2009^5,6^. Since early 2022, in the aftermath of two extreme weather events, Malawi has been experiencing its largest recorded cholera outbreak, with over 58,000 confirmed cases and 1761 deaths as of the end of May 2023^6^.

Previous genomic analysis has revealed cholera epidemics in Africa to be associated with transcontinental transmission of *V. cholerae* O1 El Tor sublineages from Asia, followed by regional cross-border spread within Africa^7,8^. To understand the origin of the current Malawi cholera outbreak, the Public Health Institute of Malawi (PHIM) partnered with the Centre for Epidemic Response and Innovation (CERI), a specialized genomics facility of the Africa Centres for Disease Control and Prevention (Africa CDC) and World Health Organization AFRO (WHO AFRO)^9^, to perform in-country genomic sequencing of *V. cholerae*. Here, using phylogenetic and phylogeographic methods combined with epidemiological modelling, we present an initial report of the genomic epidemiology of the ongoing cholera outbreak in Malawi.

## Results

On 3 March 2022, a cholera outbreak was reported following laboratory confirmation of a cholera case in Machinga district (date of symptom onset 25 February 2022). The outbreak occurred following Tropical storm Ana (28 January 2022), which caused widespread flooding and damage in southern Malawi and left >200,000 people displaced, many in emergency camps without adequate access to safe water and sanitation^6,10^. Soon after the start of the outbreak, Cyclone Gombe caused further flooding (as measured by area flooded) and heavy damage across southern Malawi, including many areas already affected by Tropical storm Ana (**Figure 1A, Figure S1**). Until August 2022, the cholera outbreak was restricted to the flood-affected areas in the southern region, when cases were reported in central and northern regions (**Figure 1A, Figure S1**). From late November 2022, during the normal rainy season, transmission intensified and cases surged particularly in Blantyre and Lilongwe, the two main urban centres (**Figure 1A**, **Figures S1 & S2**). The time varying reproduction number estimated from daily case data notably increases after 28 November before starting to decline in early January (Figure S2) and we find that flooding was positively associated with the change in the reproduction number (Materials and Methods). On 5 December 2022, the government declared a public health emergency with cases reported in all 29 districts. In the outbreak, the majority of reported cases were in young adults (<30 years) and children (**Figure S3**), whereas most deaths were in older adults (>60 years)^6^.

**Figure 1.**
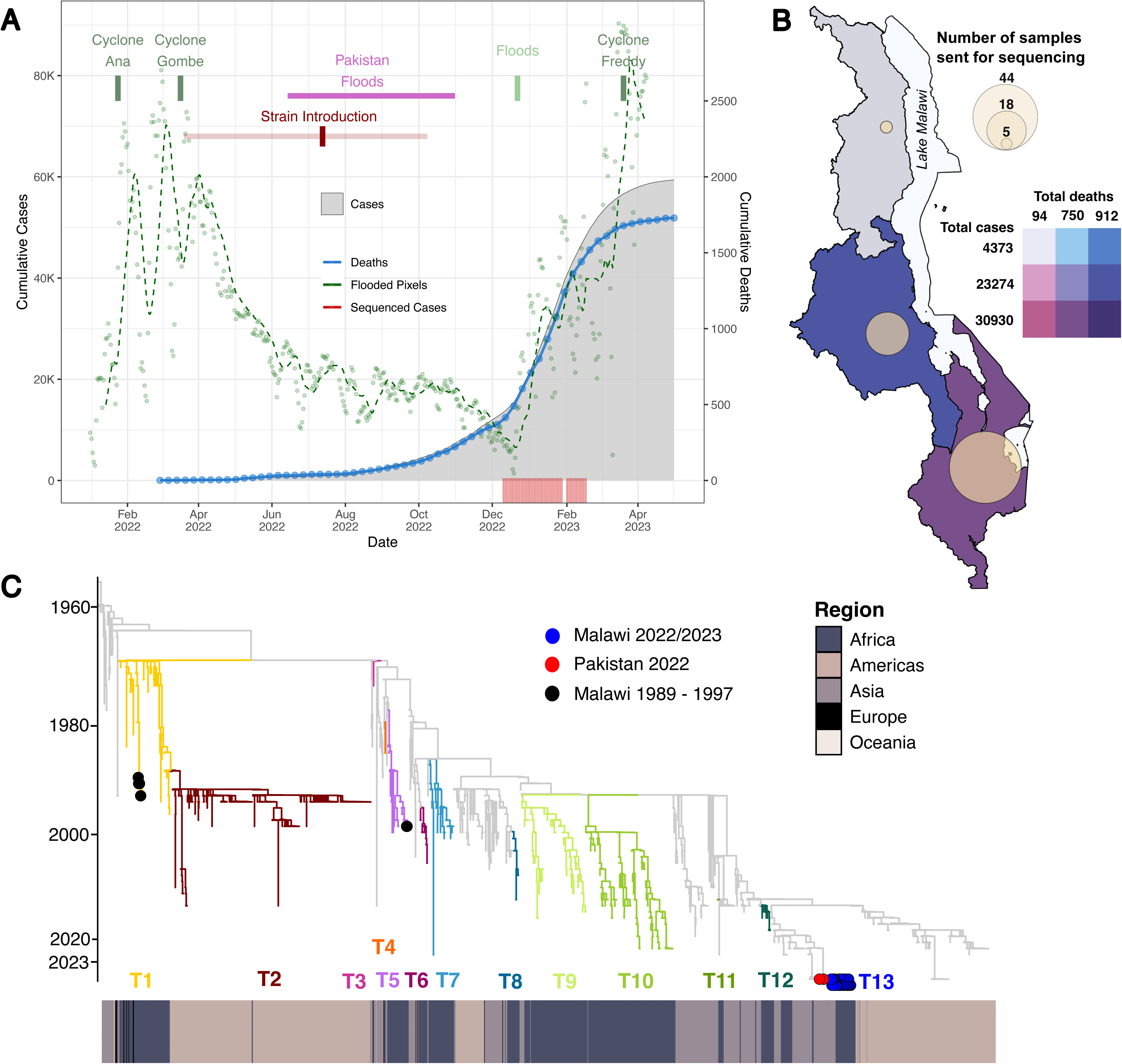

We obtained 70 *V. cholerae* isolates from stool samples collected for diagnostic culture between December 2022 and March 2023 from the southern (n=44), central (n=18) and northern (n=5) regions (**Figure 1B**). From the 70 isolates, we generated 49 high-quality, near-complete *V. cholerae* genomes (**Figure S4**). We obtained high quality SNPs (hqSNPs) from the 49 isolates by mapping to the reference wild-type El Tor N16961 strain (from the 1971 outbreak in Bangladesh)^11^. To understand the origin of this outbreak, we performed a time-scaled maximum likelihood phylogeny based on hqSNPs of the 49 genomes from Malawi and 2159 publicly available genomes from Africa (n=744), Asia (n=494), the Americas (n=899) and Europe (n=22) between 1957 and 2023 (total 2208 genomes, **Figure 1C**). The Malawian strains clustered within a well-supported monophyletic clade (bootstrap > 90%) (**Figures 1C & 2A**), closely related to isolates from the 2022 outbreak in Pakistan^12^.

**Figure 2.**
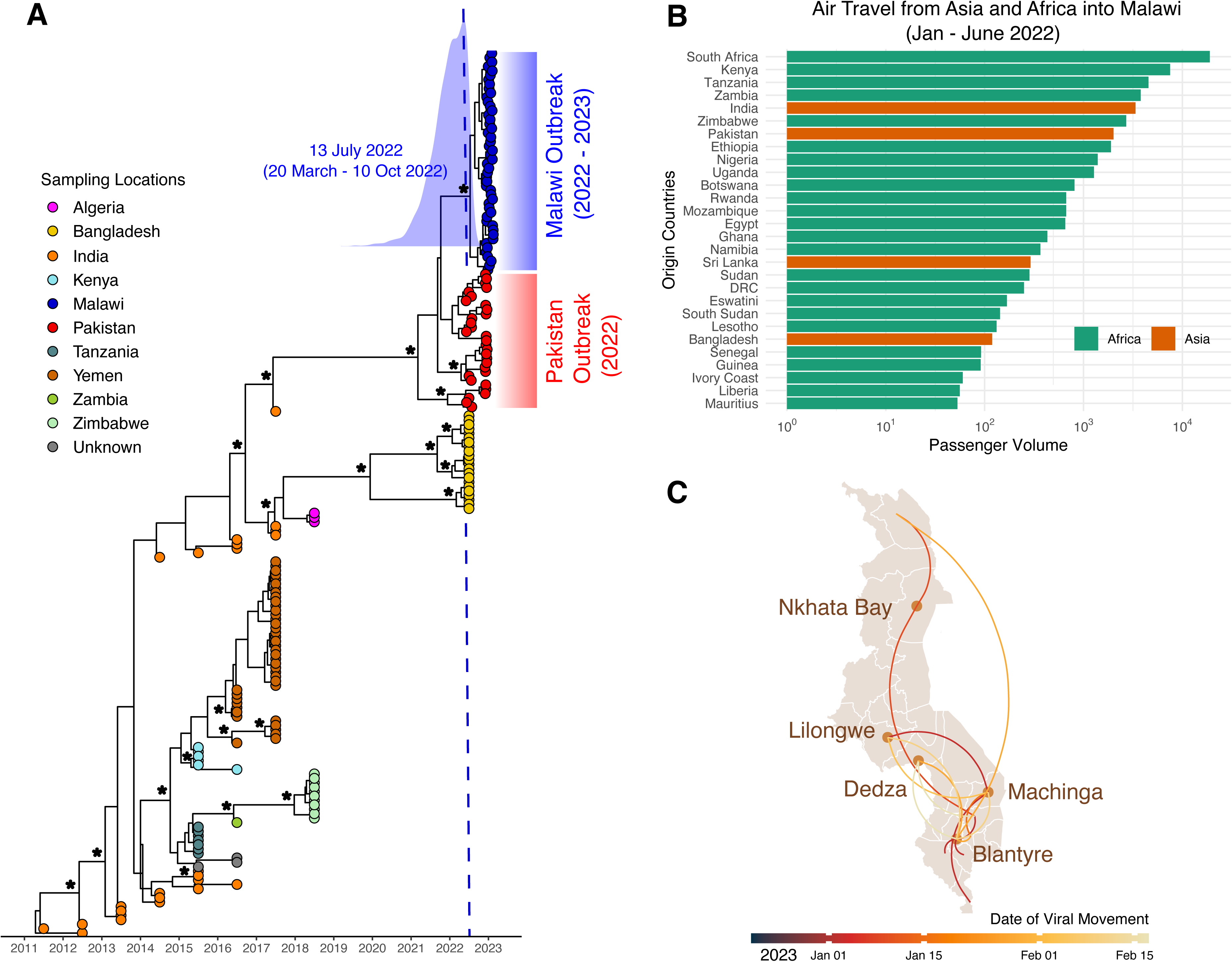

The isolates from the 2022 Pakistan outbreak clustered at the base of the Malawian clade, suggesting that the Malawi outbreak may have resulted from a long-range transmission event from Pakistan (**Figures 1C & 2A**). The time of the most recent common ancestor (MRCA) of the Malawi strains was estimated at July 2022 (95% highest posterior density (HPD) March 2022 – October 2022) (**Figure 1A**). The Pakistan cholera outbreak began in January 2022 and was exacerbated by major floods between June and October 2022 (**Figure 1A**)^13–15^. The outbreak affected the whole country and was the largest in decades, with >335 000 suspected cholera cases between 15 Jan 2022 and 15 March 2023^16^. Pakistan and Malawi are well connected in terms of human movement, with large numbers of air passengers travelling between the two during the first half of 2022 (**Figure 2B**).

Phylogeographic analysis was limited by the relatively narrow sampling date range between December 2022 and February 2023, and was thus not able to provide insights into the dispersal patterns during the early phase of the outbreak. However, the analysis suggested that in the epidemic acceleration phase, there was evidence of dispersal from multiple hubs including Machinga and Blantyre, followed by Lilongwe (**Figure 2C**).

## Conclusions

In this genomic analysis, we provide strong evidence of a link between the large cholera outbreaks in Pakistan and Malawi, consistent with prior genomic analyses revealing the importance of long-range *V. cholerae* transmission events between Asia and Africa^7,8^. Our main findings agree with another recent analysis of the Malawi outbreak^17^, and with a genomic analysis of six cases from South Africa in 2023 (three of which were epidemiologically linked to Malawi)^18^. The extreme weather events and humanitarian crises in Malawi provided the environment for the spread of *V. cholerae*, and the subsequent movement of large numbers of people may have facilitated its spread to susceptible populations in areas relatively unaffected by cholera for over a decade.

The relatively narrow sampling date range for this genomic analysis means that we cannot confidently differentiate whether the introduction of *V. cholerae* was responsible for initiating the outbreak, or if it just contributed to the later expansion of the outbreak. Additional sequencing of isolates from earlier in the outbreak may help to clarify this. We are also working with partners in other heavily affected African countries to conduct genomic sequencing of *V. cholerae* isolates, which will help with understanding the extent of cross-border regional transmission at different phases of the outbreak.

Overall, this highlights the need for coordinated global and regional cholera prevention and control efforts, and the importance of heightened awareness, data sharing and preparedness whenever outbreaks are occurring in any part of the world^19^.

## Methods

### Ethics

The CLIMADE initiative is approved by the Stellenbosch University Health Research Ethics Committee (BES-2023-24266). The sequencing of *V. cholerae* outbreak isolates in Malawi was approved by the Public Health Institute of Malawi and Malawi Ministry of Health National Health Sciences Research Committee. Isolates were anonymized and did not contain any personal identifiers.

### Epidemiology and case reporting

We utilized the WebPlotDigitizer tool (https://apps.automeris.io/wpd/) to extract data from a WHO report on cholera in the Africa region (https://extranet.who.int/iris/restricted/bitstream/handle/10665/366745/AFRO%20Cholera%20Bulletin.06.pdf). The report provided daily information on cholera cases and deaths within Malawi until April 4th 2023. WebPlotDigitizer converts information in the form of a picture to readable data in an Excel format.

### Estimation of time varying reproduction number (Rt) and relationship with flooding

We infer the instantaneous reproduction number over time, at the national level, for Malawi using a semi-mechanistic Bayesian framework described in Bhatt *et al*. ^20^. The model is estimated using the ‘epidemia’ package in R^21^, which allows us to estimate the impact of covariates such as flooding and vaccinations. The estimate of R_t is based on daily national case counts, flooding, and vaccination data. We report the mean R_t estimate, the 95% credible interval, a comparison of the observed and fitted case counts, and the effect sizes of our covariates.

We model the number of new cases, i_t’, at time t>0, through a renewal equation, and these new cases are moderated by R_t.

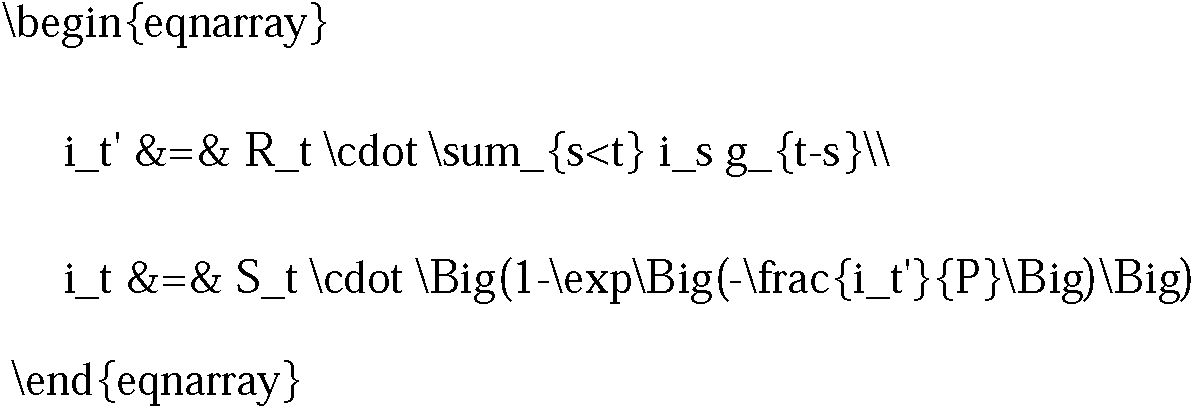

where g is the serial interval, that is the distribution of time from symptom onset of the primary case to the symptom onset of the secondary case. For cholera we assume a mean serial interval of 5 days, a standard deviation of 8 days and that g is gamma distributed with a 1-day shift^22^. S_t is the size of the susceptible population at time t and P is the size of the population and equation (2) ensures that i_t’ accounts for the depletion of susceptible individuals in the population. We give the initial susceptible population S_0 a N(0.95,0.1) prior as this provides a conservative prior which assumes that a very large majority of the population is susceptible.

We model case observations as $o_t = \sum_{s<t} i_t’\cdot \pi_{t-s}$ where π is the infection to onset distribution with a median of 1.4 days^23^.

We can form a linear predictor, consisting of fixed effects and autocorrelation terms, which is then transformed via a link function, to model the reproduction number.

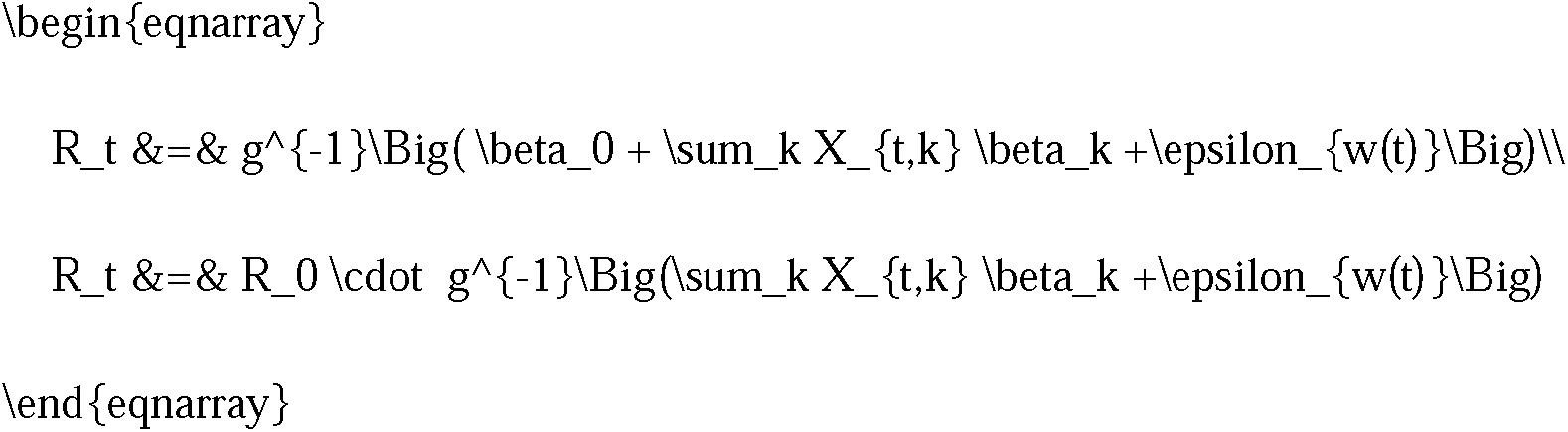

We use a *log*-link function as this ensures non-negative reproduction numbers which don’t grow exponentially. We use R_0 ∼ Normal(1.56,0.2) as a prior, where the mean is consistent with Mukandavire *et al*^24^. $\epsilon_{w(t)}$ is a weekly random walk (we use a weekly random walk rather than daily to obtain more stable R_t estimates), $X_{t,k}$ are covariates of interest and $\beta_k$ their respective effect sizes.

We use ‘epidemia’ to obtain the posterior distribution with priors as provided above. We use two covariates in our analysis:

1. **Flooding data (see details in section on flooding)** – we transform available flooding data as follows

1. 1-day difference of flooded area
2. Indicator if difference is bigger than 1000 pixels
3. Delay by 7 days to account for mean serial interval and incubation period This transformation creates an indicator time series of significant rainfall and shifts this to account for the mean serial interval and incubation period. This is a mechanistic representation for the impact of rainfall similar to (Camacho et al, 2018)^22^.
2. **Vaccinations** – two vaccination campaigns took place in Malawi against Cholera (WHO)^25^. The first vaccination campaign is not modeled as it was early in 2022. The second vaccination campaign is modeled such that we assume that the roll-out takes place over 3 months, with a delay of 10 days to account for the time lag until the vaccine is effective. We only estimate the effect size of vaccination as an intervention.

To check for robustness of our results we estimate R_t with both flooding and vaccinations, only flooding and with no covariates.

We also confirm that the estimation of R_t (without any covariates) is consistent with that of EpiEstim^26^, which is an alternative implementation of a branching process model in R. Our R_t estimates are more stable due to the weekly random walk which we use (rather than daily).

### Flooding data and processing

Much of the real or near real-time flooding data are provided by remotely sensed Earth Observation (EO) imagery. Three such are the European Sentinels^27^, the NASA MODIS satellite MCDWD product^28^, and the VIIRS satellite Flood product (NOAA&GMU Flood Version 1.0 (VNG Flood V1.0) available via the Real Earth website^29^ and associated archives^30^). The first of these tends to be focused on providing data for emergency response rather than providing long term archives; the second provides a long-term time series but stopped in 2022; and the third continues to provide ongoing daily and 5-day composites which were duly downloaded for the whole of 2022 and 2023 until April 13th. The image values distinguish ‘normal’ open water (value 99) from flood water for which values above 100 represent percentage flooding plus 100. The 5-day composite is the maximum value recorded during the period. The files are provided in simple tiled geotiff format of which two are required to cover the whole of Malawi, which must therefore be mosaicked and windowed prior to analysis.

### International passenger flight data

We evaluated travel data generated from the International Air Transport Association (IATA) to quantify passenger volumes originating from international airports and arriving in Malawi. IATA data accounts for approximately 90% of passenger travel itineraries on commercial flights, excluding transportation via unscheduled charter flights (the remainder is modelled using market intelligence).

### Sample selection, culture and DNA extraction

During the outbreak, stool samples were collected from patients presenting with cholera symptoms in district and central hospitals. Rapid diagnostic tests (RDT) were used to identify the presence of *V. cholerae* and then culture was performed on all samples which were RDT positive. Drug susceptibility testing was done on all positive cultures. All positive culture plates were taken to the National Genomic Sequencing Reference Laboratory for *V. cholerae* DNA extraction. DNA was extracted using the QIAamp DNA Mini Kit 51304 and quantified on Qubit 4 using Qubit High Sensitivity DNA kit Q32854.

For this analysis, DNA extracts were obtained from 70 *V. cholerae* isolates from samples collected between December 2022 and February 2023 from the three regions of Malawi (Southern, Central and Northern). Demographic, clinical and diagnostic data were extracted from the routine cholera surveillance system for all 70 samples.

### Sequencing

Libraries were prepared using the Illumina DNA library preparation kit and Nextera CD indexes (Illumina, San Diego, CA), according to the manufacturer’s protocol. Whole genome sequencing on all isolates was performed on the NextSeq 1000 instrument using the P2 (300) cycle kit reagents (Illumina,San Diego, CA).

### Assembly and hqSNP calling

Prior to assembly read quality was assessed with fastp^31^ and all residual adaptors were trimmed. The Snippy v4.6.0 pipeline (https://github.com/tseemann/snippy) was used to perform reference-based assembly against the N16961 strain (acc. No. NZ_CP028827.1 and NZ_CP028828.1). Variant calling thresholds FreeBayes^32^ were set as minimum 10x site coverage, minimum mapping quality 60, and minimum 90% base concordance and individual vcf files were merged using bctfools v.1.15^33,34^. Consensus genome assemblies were run through fastBaps v.1.0.8^35^ prior to recombination screening with Gubbins v.3.2.1^36^. Fasta file manipulation was performed with seqkit v2.0.0^37^ and Biostrings R package v.2.58^38^ and parsimony informative sites for phylogenetic analyses were extracted from consensus genome alignments in Mega-X v.10.0.3^39^.

In order to put our newly generated Malawian sequences into a global context we downloaded all available *V. cholerae* whole genome sequencing experiments from either the Sequence Read Archive (SRA) and/or the European Nucleotide Archive (ENA). Paired end reads were assembled and SNPs called using the exact same methodology and reference that was applied to the newly sequenced Malawian strains.

### Phylogenetic inference with cholera worldwide data set

A maximum likelihood (ML) phylogenetic tree was inferred from the parsimony informative sites using IQ-Tree^40^ to investigate the genetic relationship of the Malawian outbreak to that of other strains from around the world (sampled between 1957-2023; n=2159; Table S1). This reference set included 899 sequences from the Americas, 494 from Asia, 22 from Europe, and 744 from Africa. Due to several ongoing cholera epidemics on the African continent and the endemicity of cholera in the Indian subcontinent we ensured that our reference set also include the most recent publicly available sequences (2015 onwards) from other African countries and the Indian subcontinent (31 from Pakistan, 22 from Bangladesh, and 24 India). Phylogenetic signal was determined using a likelihood mapping test in IQ-TREE^40^. Maximum likelihood tree scaled in time was obtained with Treetime^41^.

### Phylodynamic inference

We investigated the phylogenetic relationships of our 49 new clinical strains from Malawi with 31 strains from Pakistan in 2022 (Table S1). Phylogenetic signal was determined using likelihood mapping test in IQ-TREE^40^ and temporal signal was estimated by plotting the root-to-tip divergence using TempEst^42^. The Bayesian framework was used to infer a posterior distribution of trees and estimate the time of the most common ancestor (TMRCA) of the sampled sequences. Different molecular clock models (strict or uncorrelated relaxed molecular clock) and demographic priors (constant or Bayesian Skyline Plot) were considered. We also compared different tree topologies where monophyly was enforced between the 2022-Haiti monophyletic clade and specific environmental strains sampled in 2018, in order to test hypotheses of common ancestry. Markov chain Monte Carlo (MCMC) samplers were run for 500 million generations, sampling every 50,000 generations, which was sufficient to achieve proper mixing of the Markov chain, as evaluated by effective sampling size (ESS) >200 for all parameter estimates under a given model. Hypothesis testing for best molecular clock, demographic model, and monophyly was performed by obtaining marginal likelihood estimates (MLEs) *via* path sampling (PS) and stepping-stone (SS) methods for each model to be compared, followed by the calculation of the Bayes Factor (BF), i.e., the ratio of the of the null (*H_0_*) and the alternative hypothesis (*H_A_*) MLEs^43–45^, where: *ln*BF<0 indicates support for *H_0_*; *ln*BF<2, difference barely worth mentioning; 2<*ln*BF<6, strong support for *H_A_*, and *ln*BF>6, decisive support for *H*_A_ ^46^. The best model was an uncorrelated relaxed clock with Bayesian Skyline demographic prior (Table S2). Bayesian calculations were carried out with BEAST v1.10.4 software package^45^. The maximum clade credibility (MCC) tree was obtained from the posterior distribution of trees using optimal burn-in with TreeAnnotator in the BEAST package. The MCC phylogeny was manipulated in R using the package ggtree for publishing purposes^47^.

## Data availability

Following permission from the PHIM, raw reads of the isolates that were successfully sequenced have been made publicly available and deposited at the Short Read Archive (SRA) of GenBank, Bioproject: PRJNA967700.

## Acknowledgements

CERI and the CLIMADE program are supported by grants from the Rockefeller Foundation (HTH 017), the African Society for Laboratory Medicine, the SAMRC South African mRNA Vaccine Consortium (SAMVAC), the South African Department of Science and Innovation (SA DSI).

