## Supplementary figures and images for "Genomic epidemiology of the cholera outbreak in Malawi 2022-2023"

### Supplementary Figure 1

#### Malawi Cholera Outbreak (Per District)

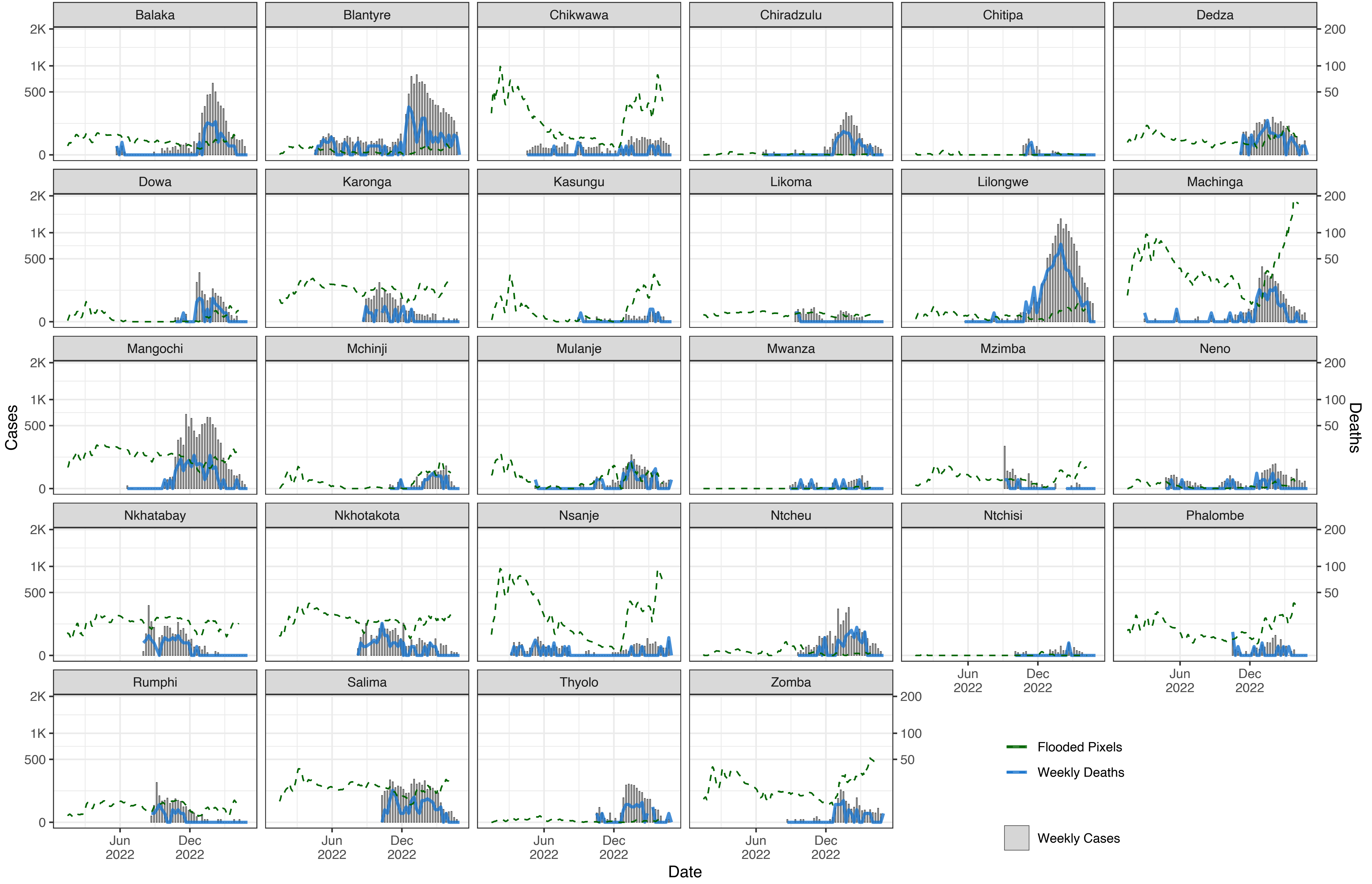

### Supplementary Figure 2

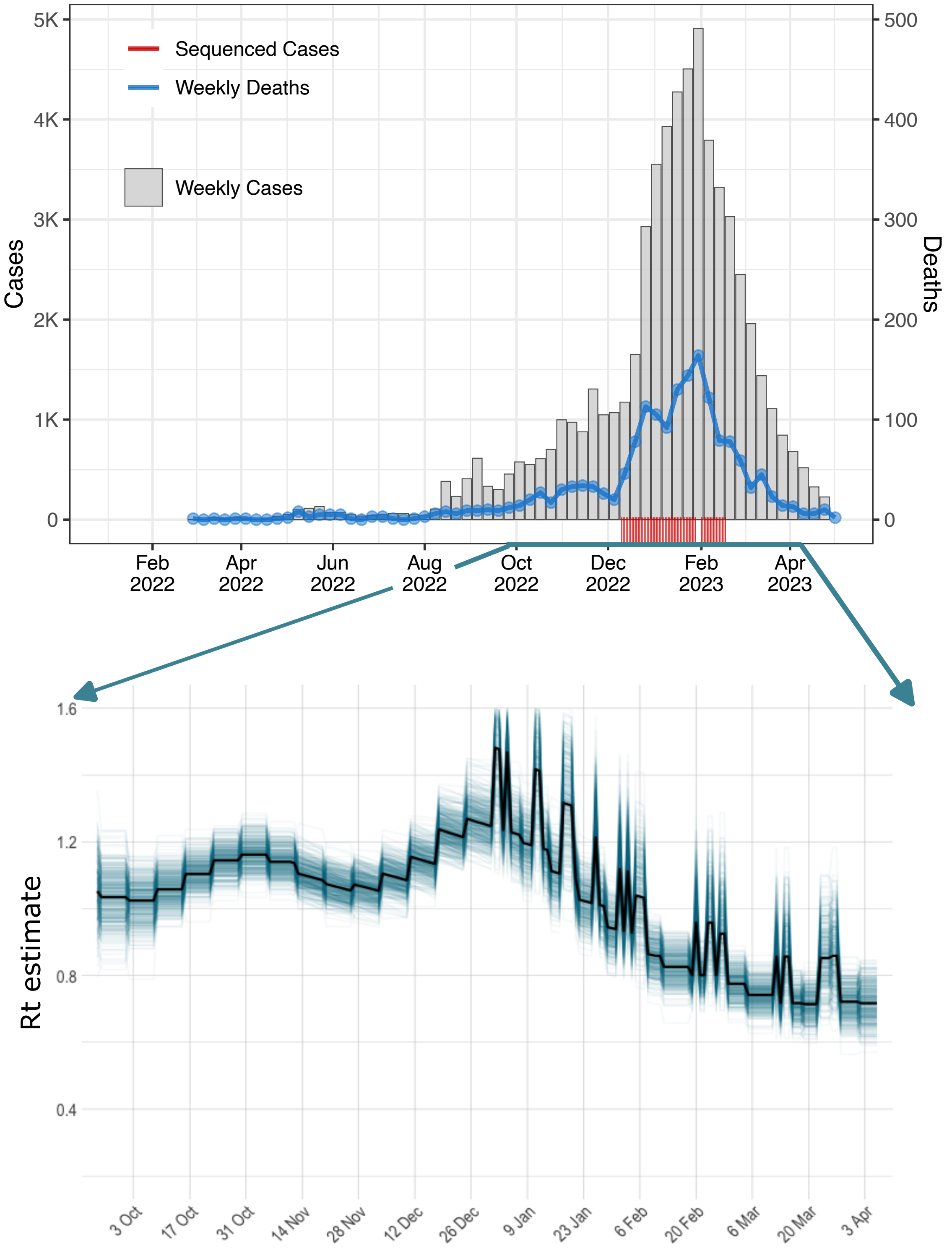

### Supplementary Figure 3

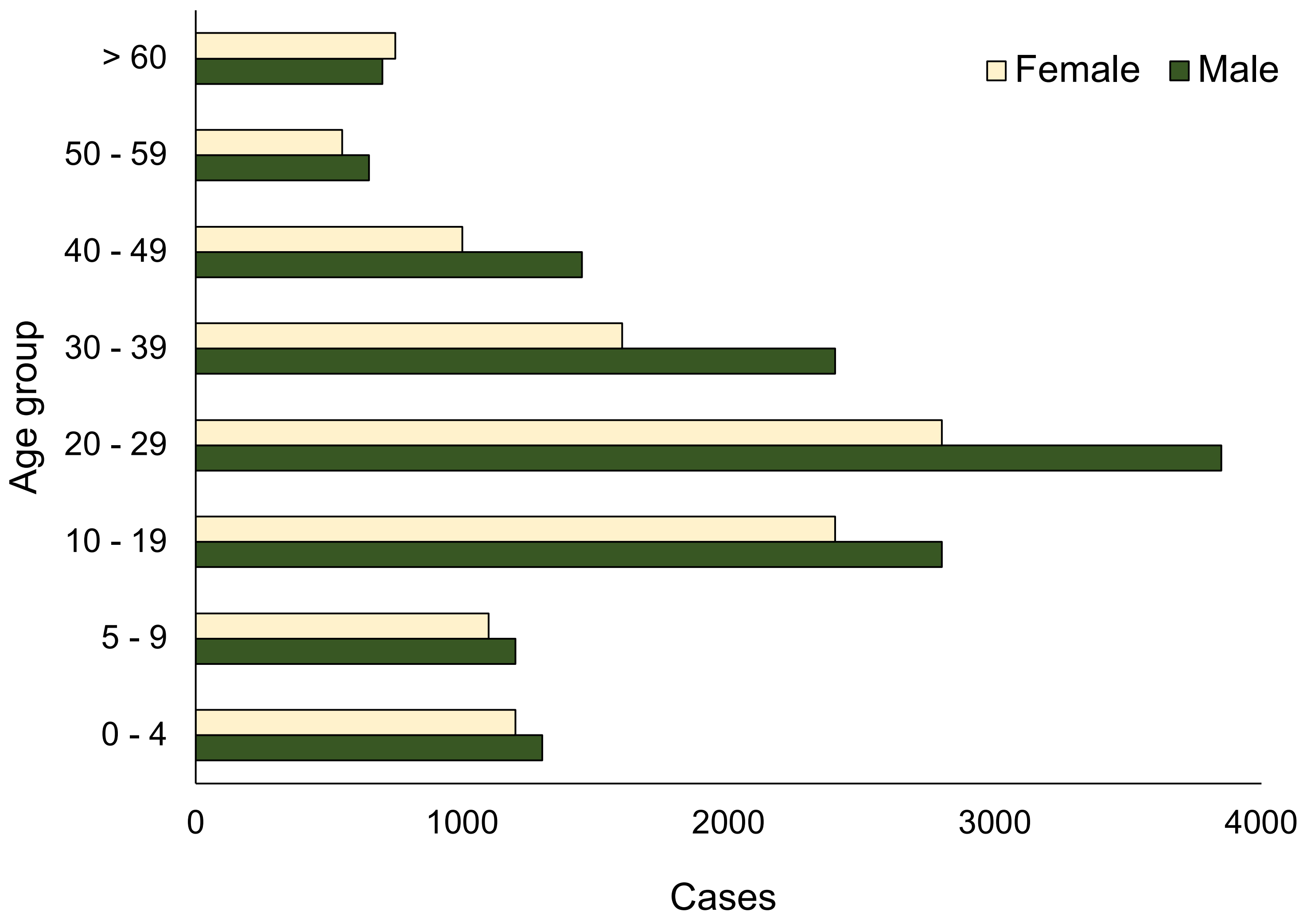

### Supplementary Figure 4

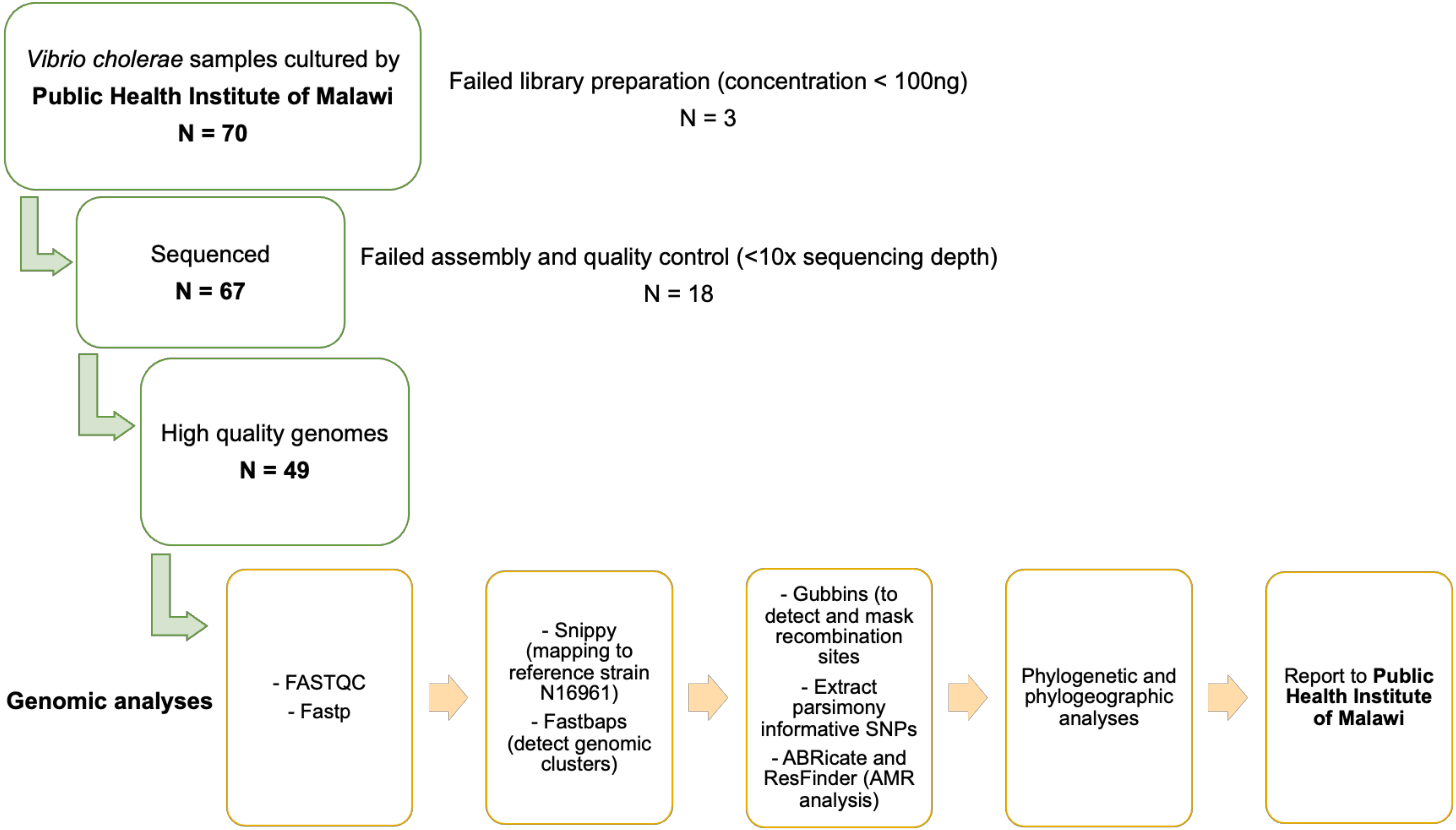
