## Supplementary Table 2 for "Genomic epidemiology of the cholera outbreak in Malawi 2022-2023"

**Table S6. Model selection of molecular clock and Bayesian demographic models to infer time-structured phylogeny.** Log of marginal likelihood estimate, ln (MLE) values obtained by Stepping Stone (SS) and Path Sampling (PS), are reported for models that either used as priors: strict (SC) or uncorrelated relaxed lognormal (UCLN) molecular clocks, and constant (CONST), non-parametric Bayesian skyline (BSP) demographic models. Bayes Factor (BF) values.

| Model | *ln*(ML)_SS_ | Ln(BF)_SS_ | *ln*(ML)_PS_ | Ln(BF)_PS_ |
| --- | --- | --- | --- | --- |
| *SC CONST* | *-2052.3* | *12.7* | *-2051.3* | *13.5* |
| *RC CONST* | *-2039.6* |  | *-2038.8* |  |
| *SC BSP* | *-2048.8* | *8.3* | *-2047.7* | *8.2* |
| *RC BSP* | -2040.5 |  | -2039.5 |  |
| *RC CONST* | *-2039.6* | *-0.9* | *-2038.8* | *-0.7* |
| *RC BSP* | *-2040.5* |  | *-2039.5* |  |
